# Detection of Peanut Allergy Sensitization and Monitoring of Type 2 Memory B cells Using Recombinant Peanut Allergen Components

**DOI:** 10.64898/2026.11.18.26368397

**Authors:** Simone Reinwald, Pei M. Aui, Rhiannon L. Fettes, Kirsten Deckert, P. Mark Hogarth, Mark Hew, Robyn E. O’Hehir, Menno C. van Zelm

**Author notes:** **Corresponding author:** Menno C. van Zelm.

## Abstract

**Background:** Peanut allergy is one of the most common causes of life-threatening food allergy, and therefore the focus of multiple clinical trials for allergen immunotherapy. Diagnosis relies on detection of allergen-specific serum IgE, which provides limited information on severity and threshold. Furthermore, objective biomarkers to predict allergic persistence and efficacy of treatment are lacking. Therefore, we aimed to incorporate recombinant peanut allergen components in a multiplex Cytometric Basophil detection assay (CytoBas) for detection of peanut sensitization, and in a memory B cell (Bmem) panel for evaluation of biomarkers of disease.

**Methods:** In 27 adult peanut-allergic patients, allergen specific IgE levels were determined by ImmunoCAP, and component specific IgE by ELISA. PBMC of all patients and of 15 non-sensitized controls were analyzed by flow cytometry with basophil markers and six fluorochrome conjugated allergen component tetramers for CytoBas, while circulating Bmem were studied including fluorescent tetramers for Ara h 1, Ara h 2 and Ara h 3.

**Results:** CytoBas demonstrated similar-to-improved diagnostic performance compared to IgE ELISAs for Ara h -1, -2, -3, -6, -8 allergens (ROCs AUC 0.79-1.0 versus 0.78-0.9). Ara h 2 discriminated all patients from controls, while combined Ara h 1, 3 and 6 (excluding major allergen Ara h 2) elicited 78-81% sensitivity with 100% specificity. Peanut-allergic patients showed expansion of the pathogenic Type 2 Bmem subset within Ara h 1-, 2- and 3-specific cells, and the Ara h 1+ and Ara h 3+ Bmem contained expanded IgG4+ and IgG1 subsets.

**Conclusions:** Peanut CytoBas displays similar to superior diagnostic accuracy to component-specific IgE serology, enabling efficient evaluation of multiple allergen components in a single test tube. Peanut-allergic individuals show increased frequencies of allergen-specific type 2 Bmem, which could be longitudinally evaluated to monitor the natural course of disease or allergen immunotherapy.

## INTRODUCTION

Peanut allergy is one of the most severe and persistent forms of IgE-mediated food allergy, carrying a significant risk of life-threatening anaphylaxis affecting 1-2% of adults and children worldwide (1). Unlike pediatric cohorts, in whom natural resolution can occur, adult peanut allergy rarely remits, presenting a lifelong disease burden (1).

Allergen specific immunotherapy (AIT) remains the only treatment that can modify the natural history of an allergic disease. Allergists are increasingly exploring AIT, particularly oral immunotherapy (OIT), for the treatment of food allergies (2, 3). To optimize the success of food specific AIT, component resolved diagnostics and biomarkers are urgently needed for precise diagnosis, selection for AIT and monitoring treatment efficacy (4).

Currently, diagnosis relies on clinical history and detection of peanut sensitization via skin prick testing and/or serum-specific IgE measurement (5). Typically, initial skin prick and serum IgE tests utilize crude peanut extract, containing a plethora of allergens, several of which are directly associated with severe responses, while others are detected by cross-reactivity (6). Among the 20 identified allergenic peanut (*Arachis hypogaea*) proteins to date, the major protein components associated with severe allergic reactions are Ara h 1 and Ara h 3 (Cupin superfamily), and Ara h 2 and Ara h 6 (prolamin superfamily) (7). Of these, Ara h 2 is the most specific marker allergen for anaphylaxis and severe allergic reactions (8), but when used in isolation has limited sensitivity (9). Other peanut allergens, Ara h 8 and Ara h 9, trigger milder responses, potentially by cross-reactivity to other plants, namely birch pollen or stone fruit (10).

The use of single allergen components in clinical testing mitigates some of the limitations of crude peanut extract, and such, molecular allergy testing is increasingly utilized (11). Despite the improved specificity of molecular allergy testing for detection of peanut allergic patients at risk for anaphylaxis, the serum IgE levels do not directly correlate with reaction severity or the dose threshold for a severe reaction (12). The current gold standard for determining threshold and reaction severity is an oral food challenge test, which carries a risk of anaphylaxis and is not routinely performed in adult peanut allergic patients (13).

Despite its utility as an accessible marker for diagnostic evaluation, soluble serum IgE does not actively mediate the allergic response. This is facilitated by IgE bound to immune effector cells, mast cells and basophils. These cells bind IgE with high affinity through FcεRI. Evaluation of allergen-specific IgE on immune cells is a strong indicator of functional relevance (14). Such measurements have shown promise with the basophil activation test (BAT), an *in vitro* assay in which fresh blood basophils are stimulated with allergen, and degranulation is assessed with flow cytometry (14, 15). While functionally relevant, the BAT requires processing of blood within 24 hours, is labor intensive and requires separate tests for each suspect allergen or component (16). Consequently, BAT predominantly remains a research tool. Our laboratory developed a pipeline for the recombinant production of allergen components to enable formation of fluorescent tetramers for use in flow cytometry (17). These allergen tetramers can bind to allergen specific IgE bound to basophils, enabling multiplex detection of functional allergen sensitization on blood basophils in a single assay (17, 18). This Cytometric Basophil assay (‘CytoBas’) has previously been evaluated against BAT, showing higher diagnostic performance and compatibility with whole blood, washed blood, as well as cryopreserved PBMCs (17). CytoBas showed >90% sensitivity and specificity for the detection of aeroallergen sensitization in a prospectively collected cohort of patients with allergic rhinitis with or without asthma (18), and here we employ it to evaluate diagnostic accuracy in peanut allergy using 6 recombinant peanut allergens in one test tube.

In parallel, recombinant allergen tetramers can be used to detect allergen-specific memory B cells (Bmem), those cells that maintain the allergic phenotype (37, 38, 39). Recently, we reported that grass pollen allergic patients have a higher frequency of a phenotypically-distinct Type 2 subset, expressing CD23, IL4Rα and germline IgE transcripts (19), which quickly differentiates into IgE secreting plasma cells upon activation (20, 21). Allergen-specific Type 2 Bmem undergo phenotypic changes during AIT for grass pollen or bee venom, including upregulation of CD29 (integrin β1) and increase proportions expressing IgG4 (22). With this Type 2 Bmem subset acting as the reservoir of pathogenic IgE in allergies, monitoring this subset can inform the clinical course of AIT.

We here developed recombinant peanut allergen components and evaluated their applications in the CytoBas assay to determine differential peanut sensitization compared to component-specific IgE serology. We further performed immunophenotyping of Type 2 Bmem in an adult population to identify biomarkers of disease and assess their potential for monitoring responses to AIT.

## METHODS

### Study participants and sample collection

Twenty-seven adults with a history of acute hypersensitivity to peanut were recruited from the Allergy Clinic, Alfred Hospital, Melbourne, Victoria, Australia (Alfred Ethics, Project 297-20) (**Suppl. Table 1**). For all patients, specificity to peanut was confirmed in childhood through skin prick testing, serum IgE (ImmunoCAP) and, when required, provocation tests. Prior to inclusion, all patients were confirmed to retain sensitization with serum-specific IgE levels of >0.35kU/L to peanut extract (ImmunoCAP^TM^, Phadia, Uppsala, Sweden). 21 of the 27 patients have been tested for pollen allergy (rye grass) in a skin prick test. Fifteen adult controls were recruited based on the absence of hypersensitivity to peanut (Monash University Human Ethics, project 2016-0289) (**Suppl. Table 2**). Lithium-heparinized blood samples were collected from all subjects and processed within 24 hours. 50µl whole blood was used for TruCount analysis (see Flow cytometry below). Plasma was collected and stored at -80°C. Peripheral blood mononuclear cells (PBMC) were isolated by Ficoll Paque (GE Healthcare, Chicago, IL) gradient centrifugation and subsequently stored in freezing media (50% fetal calf serum, 40% RPMI 1640 and 10% DMSO) in liquid N_2_ for later flow cytometric analysis (see below). The study was conducted in compliance with the Declaration of Helsinki, ethical approval was obtained (Alfred Health Human Research Ethics Committee, Project 297-20), and written informed consent was obtained prior to inclusion.

### Recombinant protein production and protein tetramerization

Recombinant peanut allergen components Ara h 1, 2, 3, 6, 8, and 9 were designed and produced, as described previously (18). Protein sequences Ara h 1.0101, Ara h 2.0201, Ara h 3.0201, Ara h 6.0101, Ara h 8.0101, Ara h 9.0101 were obtained from the Allergen Nomenclature website (allergen.org) (23). Full-length mature protein encoding constructs designed with an N-terminal Api m 1 leader sequence, a 6-His tag and Avi-tag (**Figure 1A**) (24). Codon-optimized DNA constructs for peanut allergen components were cloned into the pFastBAC vector (Thermo Fisher Scientific, Waltham, MA) before incorporation into a Bacmid for baculovirus production. Bacmids were transfected into ExpiSF9 cells and supernatant was collected after 4 days. The viral titers were determined with a flow cytometric baculovirus titering assay (25), and a new culture of ExpiSF9 was infected with a multiplicity of infection (MOI) at 0.3-0.5 and cultured at 27°C for protein expression.

**Figure 1.**
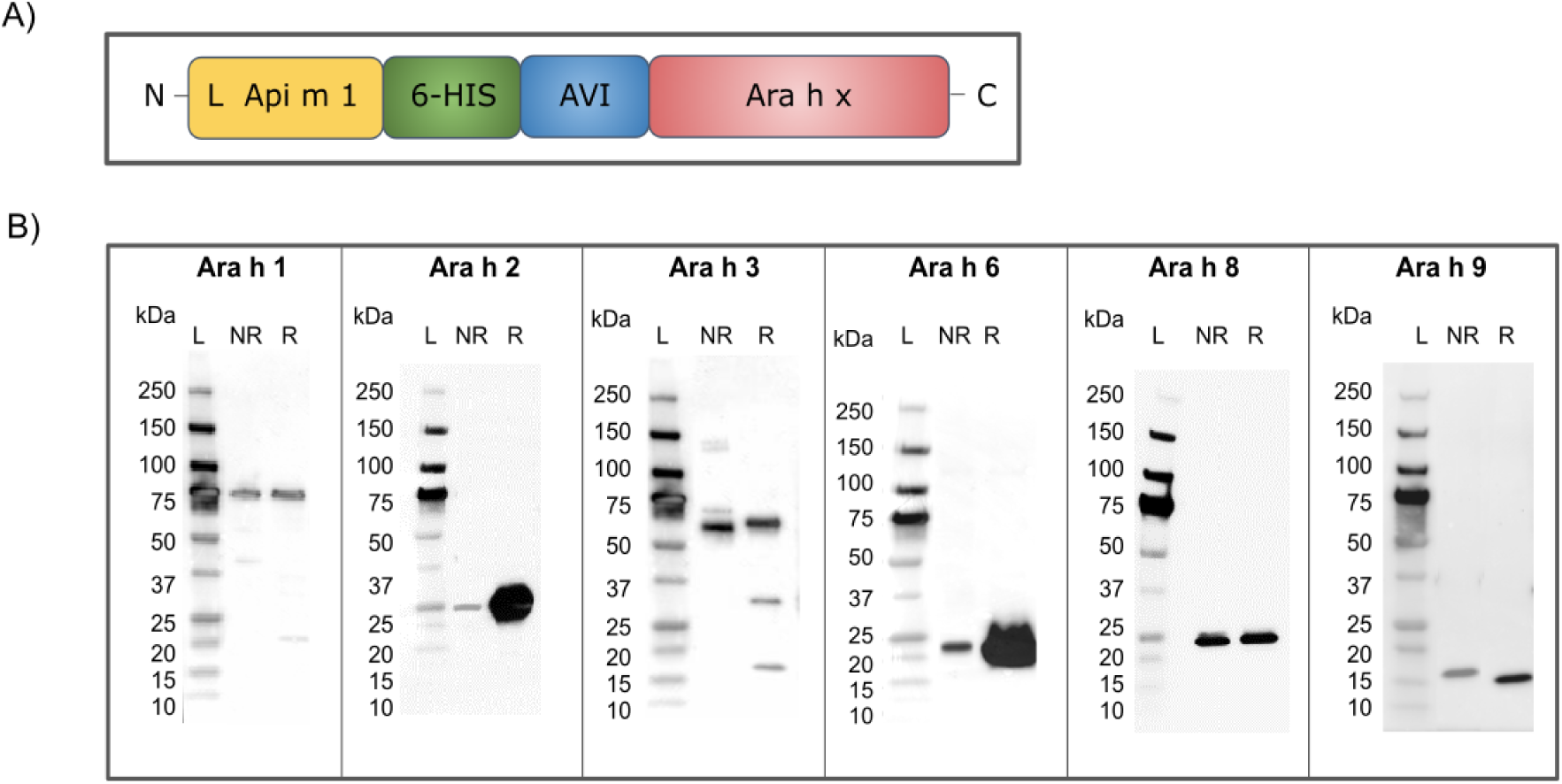
Design and recombinant production of six major peanut allergen components. (**A**) Schematic diagram of protein constructs for 6 Ara h components. Full-length mature proteins were produced with an N-terminal leader sequence for secretion, a 6-His tag and an AviTag. (B) Western Blot images of recombinant, biotinylated allergens loaded under non-reduced (NR) and reduced (R) conditions. Protein ladders (L) alongside determine the molecular weight (MW). Sequence-predicted MW: Ara h 1, 75.5 kDa; Ara h 2, 25 kDa; Ara h 3, 65.4 kDa; Ara h 6, 21.4 kDa; Ara h 8, 23.8 kDa; Ara h 9, 16.2 kDa.

All proteins were purified from culture supernatants on Cobalt columns, as described previously (18). Purified proteins were then concentrated and dialyzed in NaCl (150mM)-containing TRIS (10mM) buffer (TBS150) to enhance protein solubility (salting-in) (26). After dialysis, proteins were run under denaturing and non-denaturing conditions on a 4% to 15% mini-PROTEAN TGX Stain-Free Protein Gel (Bio-Rad Laboratories, Hercules, CA), and following blotting, evaluated with a 6x-His Tag mouse mAb (clone: HIS.H8; Thermo Fisher Scientific). The validated recombinant proteins were subjected to targeted-biotinylation incubation with a BirA enzyme (Escherichia coli 6.3.4.15; Avidity LLC, Aurora, CO, US) in SuperMix buffer (10x; Avidity LLC) at 4°C overnight (27), followed by dialysis against 10 mM TRIS (pH 8) to remove any unreacted biotin. The biotinylated proteins were tetramerized by adding fluorochrome-conjugated streptavidin, at an allergen-streptavidin molar ratio of 4:1.

### Quantification of component-specific serum IgE levels

Serum-specific IgE to single allergen components was measured by in-house enzyme-linked immunosorbent assay (ELISA) for 27 patients and 15 non-peanut sensitized controls (17, 18) (24). High-binding EIA/RIA Assay Microplates (Sigma Aldrich, Burlington, MA) were coated with recombinant peanut allergen components overnight at 4°C. Separate wells were coated with serial dilutions of recombinant human IgE-kappa (clone AbD18705; Bio-Rad, Puchheim, Germany) for generating the standard curve for interpretation of the serum IgE from samples. Plates were blocked with 5% skim milk powder in PBS and then incubated in diluted serum samples (1:20) from donors. The plate was then washed, and bound IgE was detected using polyclonal rabbit anti-hIgE (Agilent, Santa Clara, CA), which was then detected by a secondary polyclonal goat anti-rabbit HRP (Promega, Madison, WI). ELISA plates were developed with TMB solution (Life Technologies, Carlsbad, CA), and the reaction was stopped with HCl (1 N; Chem-Supply). The absorbance at OD450nm from each well was measured with the Multiskan Microplate Spectrophotometer (Thermo Fisher). Wells without any allergens were used to determine the background and were subtracted from the specific antibody IgE values.

The lower limit of detection (LOD) was determined to compare if the measured signal achieves statistical significance compared to the background noise [LOD = Mean(blanks) + 3x standard deviations (SD) across all assays run] = 2.2ng/mL IgE. Limit of quantification (LOQ) to define the lower boundary of ELISA’s quantitative range was determined [LOQ = Mean(blanks) + 10 x SD (blank)] = 4.307ng/mL IgE.

### Flow cytometry

#### Trucount

Absolute numbers of B cells, T cells, NK cells and monocytes were determined in whole blood from each sample, as described previously, (24) with the Trucount Absolute Counting tubes (BD Biosciences, Franklin Lakes, NJ). Briefly, fresh blood samples (50μL) were stained with the antibody cocktail mix containing CD3, CD4, CD8, CD16, CD19, CD45 and CD56 (**Suppl. Table 3**) in the dark for 15 mins. Staining was stopped by adding 500μL of BD Lysis solution (1x) to the tube, followed by analysis on the FACSLyric or LSRII (both BD Biosciences). The absolute count of each leukocyte and lymphocyte subpopulation was calculated as per the manufacturer’s instructions.

#### CytoBas

Individual vials of 1mL cryopreserved PBMC (10 million) were thawed in a 37°C water bath until crystal arose, followed by the addition of 1ml FCS and 8ml of RPMI-1640 (Sigma Aldrich). Samples were washed again with FACS buffer (0.7% BSA and 0.1% sodium azide in PBS) and then strained through a 70 μm filter for cell count and viability determined with the automated cell counter (Nexcelom, Lawrence, MA). 6-9 million cells of PBMC were incubated with the six peanut allergen tetramers (1μg each per 100μL stain) in the dark for 5 minutes at room temperature. In a separate tube, the remaining 2 million PBMC incubated with equal amounts of streptavidin-only conjugates (BUV395, BUV615, BUV737, BV480, BV711, and PE) (24, 28). Both sample tubes were then incubated with the backbone antibodies CD45-PerCP-Cy5.5 (2D1; BD Biosciences), CD123-cFluorBYG781 (SK3; Cytek BioSciences, Fremont, CA), IgE–fluorescein isothiocyanate (polyclonal; Thermo Fisher Scientific), HLA-DR–allophycocyanin (APC) (L243; BioLegend, San Diego, CA) and viability dye viaDye Red (Cytek BioSciences) (**Suppl. Table 4**) in the dark for 15 minutes at room temperature. Subsequently, cells were washed with FACS buffer and fixed with 2% paraformaldehyde for 20 minutes at room temperature. After staining and fixing, cells were washed twice with 2mL FACS buffer, then resuspended in FACS buffer to run on a five-laser Cytek Aurora (Cytek Biosciences). The standard Cytek Assay Settings were employed and were adjusted daily utilizing the SpectroFlo Quality Control beads, according to the manufacturer’s recommendations (29). Samples were acquired using the live unmixing function on SpectroFlo and were run at a flow rate of 3500-5500 events per second. PBMC viability ranged from 75-95% and this was >95% for basophils. Basophils comprised 0.2-0.35% of the mononuclear cell population after thawing, and >5000 basophil events were acquired per analysis to obtain reliable median fluorescence intensities for allergen-tetramer binding to the basophil population.

#### Allergen-specific memory B cells

Cryopreserved PBMC were thawed and washed with FACS buffer (0.7% BSA and 0.1 % sodium azide in PBS) and then strained through a 70 μm filter. Cell count and viability were determined with an automated cell counter (Nexcelom, Lawrence). 10-15 million PBMC were incubated with 5 μg/ml of each of the Ara h 1 and Ara h 2 tetramers (n=27 donor samples) or Ara h 3 tetramers (n=18 donor samples) for 3 mins, followed by incubation with a cocktail of antibodies: CD3, CD19, CD21, CD23, CD27, CD29, CD38, CD45, CD124 (IL-4Rα), IgA, IgD, IgE, IgG_1_, IgG_2_, IgG_3_, IgG_4_, IgM, and with Live dead Blue (**Suppl. Table 5**), in a total volume of 250μL FACS buffer and 10% brilliant stain buffer for 15 mins at RT. In parallel, ∼5 million PBMC were incubated with strep controls of BUV395-, BV421-, BV480-, BUV737-, BUV496-or BV650-strep with the same procedure, followed by incubation with a minimal antibody backbone, including CD3, CD19, CD38, CD27, IgE and Live dead Blue. After staining, cells were washed with FACS buffer and then fixed with 2% paraformaldehyde for 20 minutes at RT. Samples were then washed again and resuspended in FACS buffer for analysis on a 5-laser Cytek Aurora (Cytek Biosciences). Standard Cytek Assay Settings, which were adjusted daily utilizing the SpectroFlo Quality Control beads, were used according to the manufacturer’s recommendations.(29) Light scatter settings were fine-tuned for lymphocyte identification. The live unmixing function on SpectroFlo was employed, and samples were run at a flow rate of ∼5000 events per second.

Total Bmem and Ara h 1, Ara h 2, and Ara h 3-specific subsets were immunophenotyped (**Suppl. Figures 3 and 4**). In brief, within CD19^+^ B cells, CD38^dim^ mature B cell subset was defined, subsequently Bmem were defined by excluding IgD^+^CD27^-^ naive B cells. Allergen-specific Bmem were defined by double discrimination with Ara h 1, Ara h 2, or Ara h 3 tetramers (30). Within the total and allergen-specific Bmem populations, Ig isotypes and Ig subclasses were identified with the corresponding antibodies. Gating was performed on total B cells, and the same gates for subsetting were then applied to antigen-specific cells. Using the absolute B-cell counts obtained from the TruCount analysis, B-cell subset frequencies were converted into cells per microliter of blood.

### Data Analysis and Statistics

Statistical analyses of flow cytometric data were conducted with GraphPad Prism (v9.0.1; GraphPad Software, Dotmatics, Boston, MA). The nonparametric Mann-Whitney U-test was used for non-paired data analysis. Receiver operating characteristic (ROC) curves were created to determine the performance capabilities (specificity and sensitivity) of allergen components in serology and CytoBas assays between sensitized patients and controls. The nonparametric Delong test was used to compare the area under the curve (AUC) values obtained from ROC analysis between the CytoBas and serology tests. For all tests, p<0.05 was considered statistically significant. Diagnostic performance was evaluated using a 2 x 2 contingency table to classify outcomes as true positive, false positive, true negative, and false negative relative to the reference standard. Test specificity was calculated as (TN) / (TN + FP).

## RESULTS

### Detection of peanut allergen component sensitization with flow cytometric staining of basophils: Peanut Differentiation CytoBas

Six peanut allergen components, Ara h 1, -2, -3, -6, -8 and -9, were generated as recombinant proteins in a Baculovirus expression system with the Api m 1 leader sequence, a 6-His tag and an Avi-tag (**Figure 1A**), and validated on stain-free Protein gel and anti-His Western Blot analysis (**Figure 1B**). Each allergen component was tetramerized with a distinct fluorochrome□conjugated streptavidin for flow cytometric detection of allergen sensitization on basophils (CytoBas) in combination with 4 fluorochrome□conjugated antibodies routinely used to identify basophils (CD45, CD123, IgE, and HLA□DR) and a viability dye to exclude dead cells. Markers were chosen for reliable detection of basophils, and the fluorochromes to minimize spectral overlap. A second staining was performed on each sample containing the same antibodies and fluorochrome□conjugated streptavidins without proteins as the negative control. Using a 5□laser spectral flow cytometer, the 11 fluorescent spectra were clearly resolved with minimal spectral overlap, and the panel reliably identified basophils (**Figure 2A**). Within the basophil populations, the MFI of each tetramer was visualized together with strep-only staining for that patient, and the ratio of MFI of the tetramer over the strep□only control was defined (**Figure 2B**). Sensitization profiles vary per individual (**Figure 2B**). Following application of a cut-off for MFI ratio of 2, as defined in to previous CytoBas studies for aeroallergen and bee venom components (17, 18), none of the 15 controls showed positivity for the anaphylaxis-associated components Ara h 1, 2, 3 and 6, and 2/15 were positive for Ara h 8 and 6/15 for Ara h 9 (**Figure 2C**). Ara h 2 was most discriminatory with all 25/25 patients showing positivity, followed by the other anaphylaxis-associated components Ara h 6 (19/25), Ara h 3 (14/18) and Ara h 1 (8/18). The majority of peanut sensitized patients showed positivity for Ara h 8 (15/25) and Ara h 9 (17/25) (**Suppl. Table 6**).

**Figure 2:**
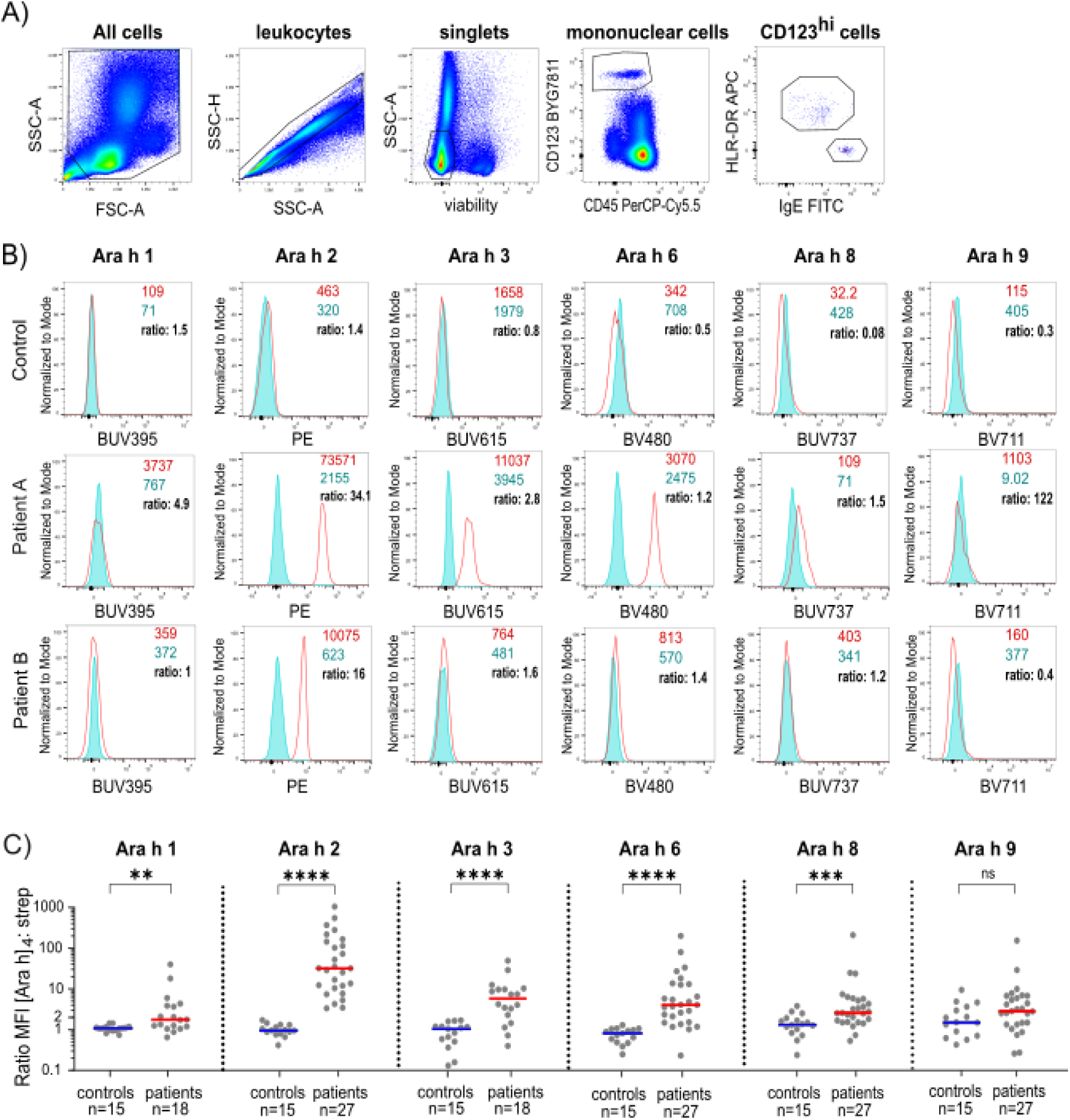
Multiplex CytoBas panel for detection of sensitization to 6 peanut allergen components on basophils. **(A**) Gating strategy for selection of basophils within cryopreserved PBMC samples. Basophils were defined as CD45^dim^ CD123^+^ HLA-DR^-^ IgE^+^ (**B**) Histograms depicting overlays of the streptavidin-only control (strep; blue) with the allergen component tetramer staining on basophils from one non-sensitized control and two peanut-sensitized individuals to display individual sensitization patterns. Each plot displays the median fluorescence intensities (MFI) and ratio of allergen staining over strep control. (C) Staining ratios for all 6 peanut components determined with a single multiple CytoBas tube of allergic (red) and non-peanut sensitized individuals (blue). Statistics: Mann-Whitney U test, * p ≤ 0.05, ** p ≤ 0.01, ***p ≤ 0.001, **** p ≤ 0.000, ns = non-significant.

### Diagnostic capacity of Peanut Differentiation CytoBas

To omit the application of an arbitrary cut-off value, the diagnostic performance of PeanutDiff CytoBas was evaluated using Receiver Operating Characteristic (ROC) analysis of patients and controls for each of the 6 components (**Figure 3A**). The area-under-the-curve (AUC) was greatest for Ara h 2 (1.0), followed by the other anaphylaxis-associated components: Ara h 6 (0.98), Ara h 3 (0.89) and Ara h 1 (0.79). The AUCs were lower for Ara h 8 (0.80) and Ara h 9 (0.64).

**Figure 3.**
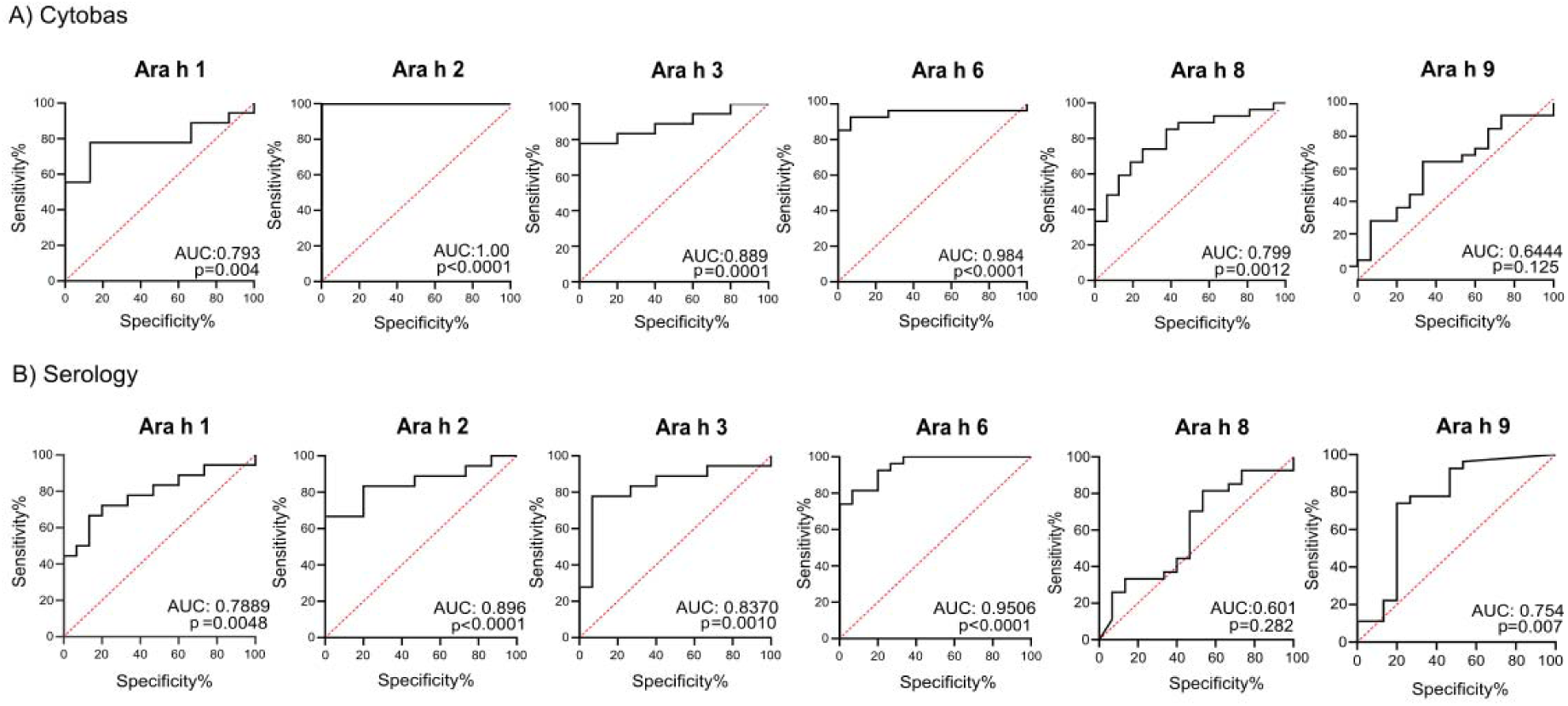
Evaluation of diagnostic performance of CytoBas and IgE serology for each peanut allergen component. **A**) ROC analysis based on staining intensity as determined by CytoBas of [Ara h 1]_4_ –BUV395, [Ara h 2]_4_ -PE, [Ara h 3]_4_ –BUV615, [Ara h 6]_4_ –BV480, [Ara h 8]_4_ –BUV737, [Ara h 9] _4_–BV711. **B**) ROC analysis based on serum peanut-specific IgE to 6 Ara h components. Included are 15 non-peanut sensitized controls, and either 18 (for Ara h 1, 3) or 27 peanut sensitized patients. The areas under the curve (AUC) and significance are depicted (Delong test). The p-value for the calculated AUC was determined using the non-parametric Mann-Whitney U test.

To compare the diagnostic capacity of PeanutDiff CytoBas with state-of-art serum specific IgE ELISA, the same 6 allergen components were used for detection of serum IgE in all 27 patients and 15 controls (**Suppl. Figure 1**). This yielded AUCs ranging from 0.60-0.89. Notably, serum IgE to Ara h 2 was less accurate than CytoBas (AUC= 0.89 vs 1.0; *p*= 0.0001), and serum IgE to Ara h 1, 3 and 6 displayed slightly lower AUC values than CytoBas (**Figure 3**). Correlation of serum IgE versus component-specific CytoBas revealed a weak association between the ranks for each of the allergens used (**Suppl. Figure 2**).

Overall, CytoBas with Ara h 2 performed the best in separating patients from controls in our cohort. However, not all anaphylactic patients show sensitization to Ara h 2 (31). As our PeanutDiff CytoBas incorporates three other anaphylaxis-associated components, we evaluated whether combined analysis of Ara h 1, 3 and 6 has a high diagnostic performance. Excluding Ara h 2, combined evaluation showed 100% specificity and 78-81% sensitivity; all patients in our cohort were positive for at least one of these 3 components (**Figure 4, Suppl. Table 6**). The PeanutDiff CytoBas provides a sensitive and specific method to detect peanut allergen component sensitization, with positivity for Ara h 1, 2, 3 and/or 6 being indicative of severe responses to peanut.

**Figure 4.**
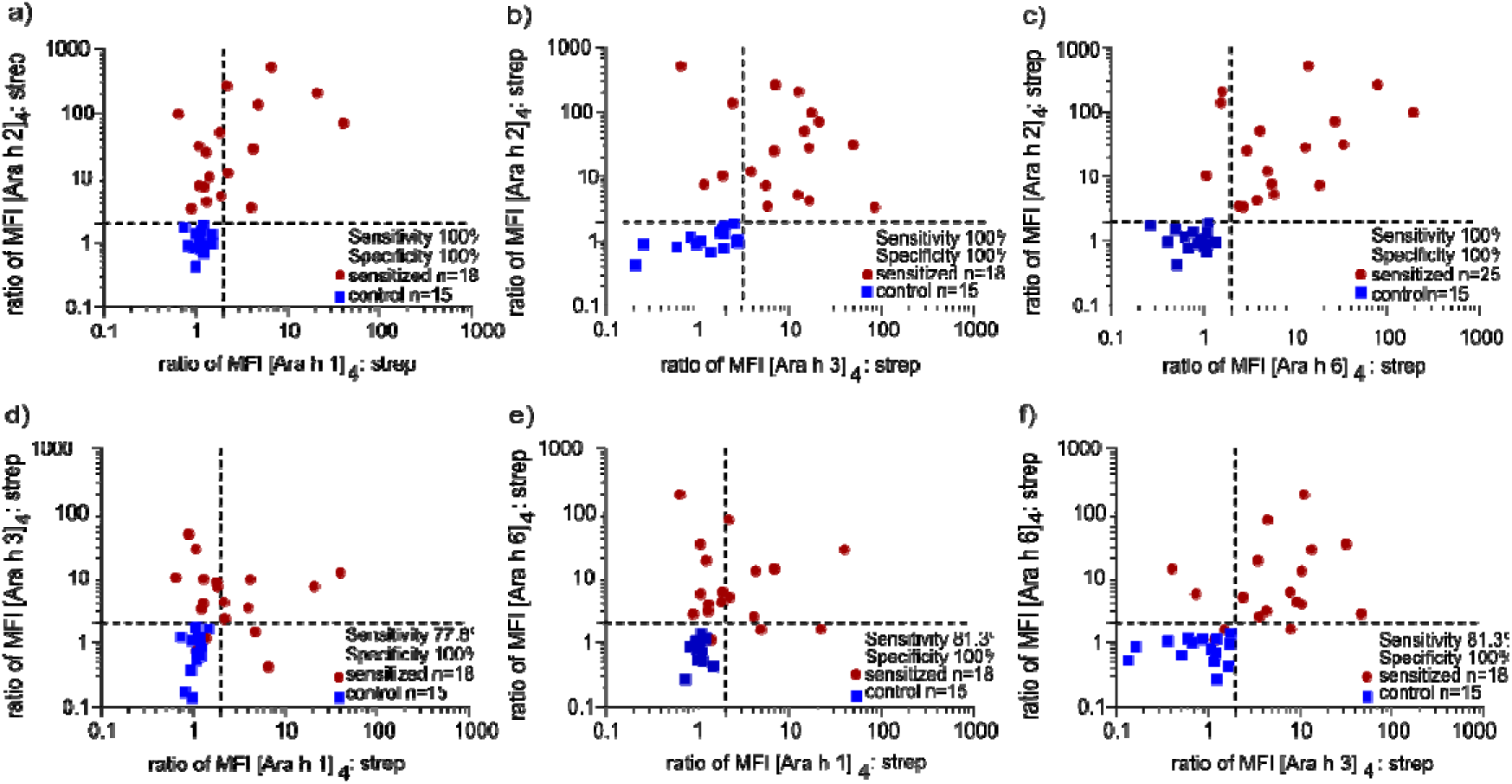
Combined diagnostic performance of Ara h 1, 2, 3 and 6 with Peanut Differentiation CytoBas for detection of allergen sensitization. Scatterplots depicting MFI ratio of allergen: Streptavidin of [Ara h 2] 4 – PE versus a) Ara h 1 BUV395, b) [Ara h 3] 4 – BUV615 and c) [Ara h 6] 4-BV480. Performance shown in d) Streptavidin of [Ara h 3] 4 - BUV615 versus [Ara h 1] 4 –BUV395 allergen, e) Streptavidin of [Ara h 6] 4 -BV480 versus [Ara h 1] 4 –BUV395 and f) [Ara h 3] 4-BUV615 on basophils from peanut sensitized and control donors. The depicted sensitivity and specificity values are based on positivity of either or both components. Cut-off values at the ratio of 2 (depicted by dotted lines) were selected in accordance with established technical performance baselines for these diagnostic modalities (17, 18).

### Immunophenotype of peanut component specific memory B cells

At the cellular level, peanut allergen-specific Type 2 memory B (Bmem) cells sustain the allergic phenotype, and may serve as biomarkers of treatment efficacy, given their rapid phenotypic transition following allergen immunotherapy (AIT) (22). Therefore, we developed a B-cell flow cytometry panel for peanut allergy, incorporating Ara h 1, Ara h 2 and Ara h 3 tetramers, each on two distinct fluorochromes, and 17 markers plus a live-dead dye. We immunophenotyped the B-cell compartment in all 27 patients (analyzing Ara h 1 and Ara h 2-specific memory) and in 18 patients Ara h 3-specific memory, plus 15 controls. Total Bmem counts ranged from 20-270 cells per µL and were not significantly different between patients and controls (**Figure 5A**). Peanut allergen-specific Bmem were well-detectable, although very infrequent (0.02-0.6% of B cells in blood), similar to our previous observations for grass pollen and bee venom (19, 32, 33). Ara h 1-, Ara h 2- and Ara h 3-specific Bmem counts were significantly higher in patients than in controls (**Figure 5A**).

**Figure 5.**
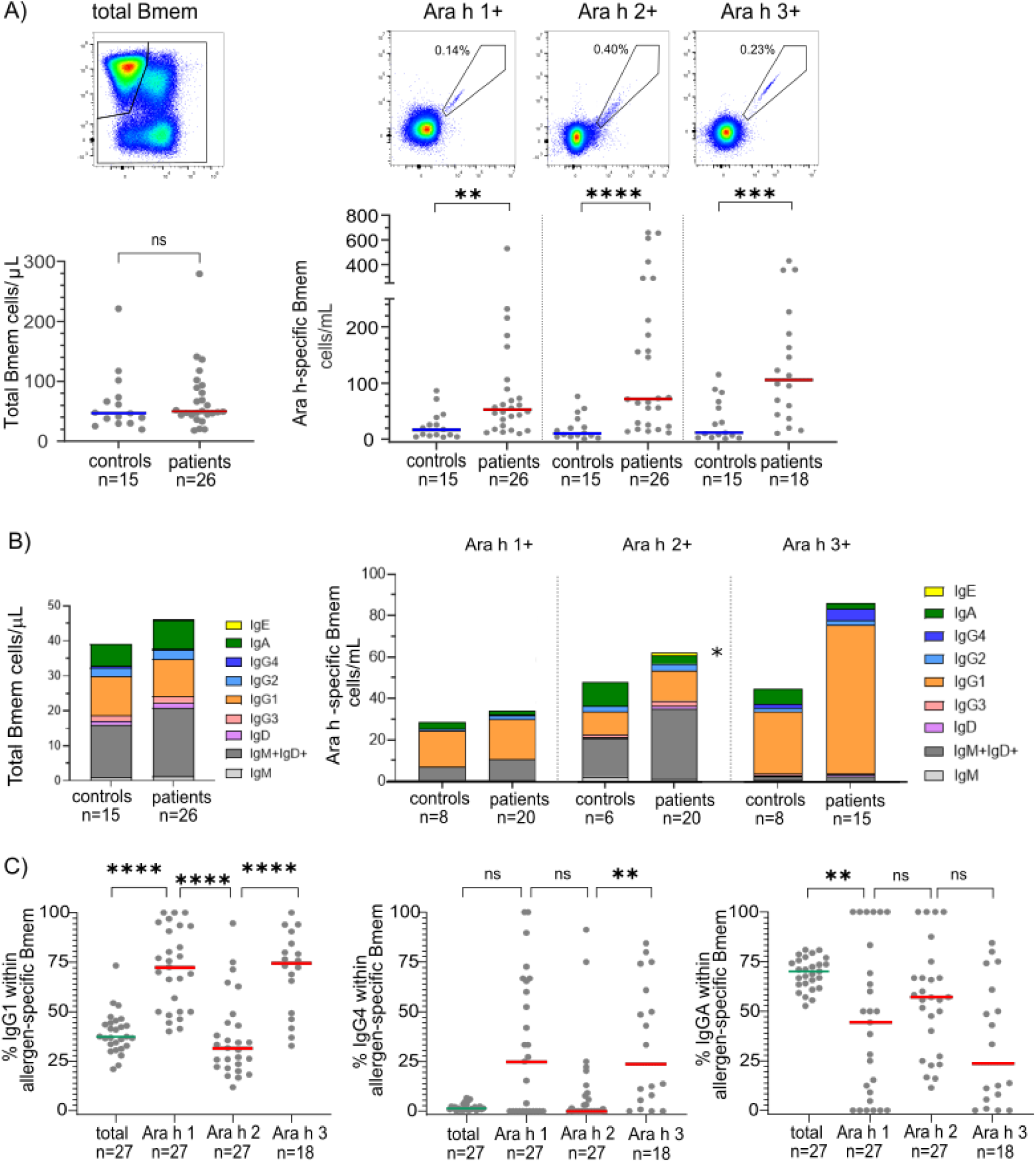
Immunophenotypes of total, Ara h 1, Ara h 2 and Ara h 3-specific Bmem. (**A**) Absolute numbers of total Bmem, and Ara h 1, Ara h 2 and Ara h 3 specific Bmem in blood of controls (n=15) and peanut-sensitized patients (n=26). The median of controls is indicated as blue line, for peanut allergic donors as red lines. (**B**) Median cell numbers of total Bmem (left) and Ara h-specific Bmem (right) expressing IgM-only, IgM+IgD+, IgD-only, IgG3, IgG1, IgG2, IgG4, IgA and IgE for non-sensitized controls (n=15) and peanut-sensitized patients (n=26). The numbers of individuals per category are indicated underneath, only samples that contained >30 events in the Bmem gate were included. (**C**) Percentage of total, Ara h 1, Ara h 2 and Ara h 3-specific Bmem expressing IgG1, IgG4 and IgA for non-sensitized controls (n=15) and peanut-sensitized patients (n=27, for Ara h 3-specific Bmem n=18). Statistics: Mann-Whitney U test, * p ≤ 0.05, ** p ≤ 0.01, ***p ≤ 0.001, **** p ≤ 0.0001, ns = non-significant.

Within total and allergen-specific Bmem, subsets were defined based on distinct Ig isotype and IgG subclass expression (**Suppl. Figure 3**). The total Bmem compartments of patients and controls consisted of three dominant populations being either IgM+IgD+, IgA+ or IgG1+, and smaller subsets expressing IgG3, IgG2 or IgG4. No significant difference between patients and controls was observed (**Figure 5B**). Within Ara h 1, Ara h 2 specific Bmem, the dominant populations were IgG1+ and IgM+IgD+, whereas in Ara h 3 specific Bmem IgG1 was the most dominant subset (**Figure 5 B, C**). In addition, IgA subset expression is reduced in Arah 2, Ara h 3 and to a smaller extent Ara h 1-specifc Bmem, but did not reach significance (**Figure 5 B, C**). Significantly more Ara h 2 specific Bmem of patients expressed IgE (p<0.05) (**Figure 5B**).

Type 2 Bmem were defined on the basis of surface expression of both CD23 and CD124 (IL-4Rα) (**Suppl. Figure 4**) (20, 21), and this subset was more frequent with a higher absolute cell count within total Bmem of patients than controls (p<0.001) (**Figure 6A**). Similarly, Ara h 1, -2 and -3 specific type 2 Bmem were more frequent in patients than in controls (all p<0.001). Within total type 2 Bmem, patients had a smaller fraction expressing IgG4 than controls. Within Ara h 1, -2 and -3 specific type 2 Bmem, IgG4 expression was detected in a minority of patients. No differences were seen with controls, although only a few controls had sufficient allergen-specific Type 2 Bmem for reliable detection of IgG4 (**Figure 6B**). CD29 expression was not different between controls and patients for total and antigen-specific Type 2 Bmem. Similar to IgG4 analysis, only a few controls displayed sufficient allergen-specific Type 2 Bmem (**Figure 6C**).

**Figure 6.**
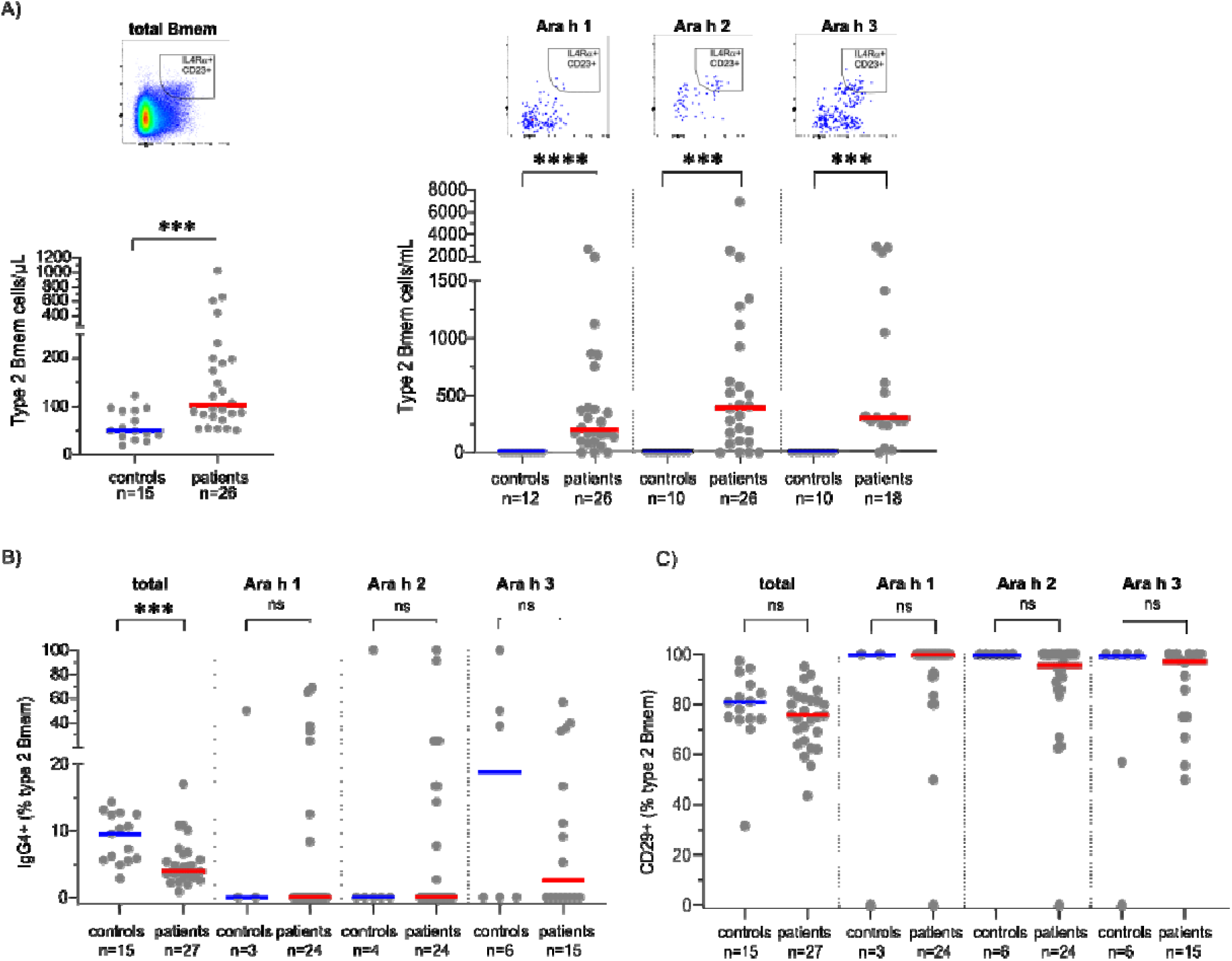
Type 2 Bmem numbers within total and allergen-specific Bmem and surface marker expression of IgG4 and CD29. (**A**) Absolute numbers of Type 2 Bmem were defined as IL4R+ and CD23+ cells within total, Ara h 1-, Ara h 2- and Ara h 3-specific Bmem. (**B**) Frequency of IgG4 expressing cells with the total, Ara h 1-, Ara h -2 and Ara h 3-specific type 2 Bmem. (**C**) Frequency of CD29 expressing cells within total, Ara h 1-, Ara h 2- and Ara h 3-specific type 2 Bmem. (**B** and **C**) Only data from samples with more than 30 events within the type 2 Bmem population were included in the analysis to ensure that the calculated correlation reflected true biological phenotypes rather than stochastic variation in low-abundance samples. Statistics: Mann-Whitney U test, * p ≤ 0.05, ** p ≤ 0.01, ***p ≤ 0.001, ns = non-significant.

Overall, we confirm and extend previous observations that the Type 2 Bmem phenotype is more frequent in allergic donors than in non-allergic controls. Peanut allergen component Type 2 Bmem are readily detectable and represent strong candidate biomarkers for allergen-specific treatment evaluation.

## DISCUSSION

In this study, we demonstrated the capacity of flow cytometric evaluation of immune cells in blood with recombinant allergen components for precise diagnostic profiling and the potential to evaluate the pathogenic B-cell reservoirs in adult peanut allergy.

A single flow cytometric assay for the simultaneous detection of sensitization to peanut allergen components on basophils showed 100% sensitivity and specificity using four anaphylaxis-associated components. In addition, a B-cell panel incorporating three allergen components enabled reliable detection and immunophenotyping of pathogenic Type 2 Bmem.

We here present the design and application of a single CytoBas panel for the evaluation of peanut allergen sensitization using six major allergen components. The cytometric detection of peanut allergen component binding to basophils demonstrated diagnostic performance comparable to, or exceeding, that of IgE serology determined by ELISA for 5 out of 6 components (Ara h 2, Ara h 3, Ara h 6 and Ara h 8, but not Ara h 1). The analyzed cohort comprised adult patients, each with a history of severe allergic reactivity and confirmed sensitization to Ara h 2, yielding 100% sensitivity and specificity. Previous studies have demonstrated that sIgE to Ara h 2 has a higher positive predictive value for clinical reactivity than either whole-peanut sIgE or skin prick testing (SPT) alone (34). However, not all patients with severe reactions show Ara h 2 sensitization, resulting in limited sensitivity of single component analysis (9). Importantly, combined evaluation of Ara h 1, 3 and 6 in our cohort showed 100% sensitivity and specificity as well, demonstrating the capacity of this multiple analysis to detect relevant sensitization when Ara h 2 is negative. Finally, inclusion of Ara h 8 and Ara h 9 can inform on cross-reactivity and mild responses in those patients without sensitization to Ara h 1, 2, 3 and 6.

The component Ara h 1 displayed lower diagnostic performance than Ara h 2 in CytoBas and ELISA, with a very comparable AUCs between assays (**Figure 3**). This may indicate that most in this patient cohort are not sensitized to Ara h 1. Other contributory factors could include protein quality, folding and tetramer formation. All recombinant allergen components were produced by the same method and were confirmed to be successfully biotinylated by Western blot analysis prior to tetramer assembly (**Figure 1B**). To minimize inter-assay variability in all our analyses, we used a single production batch of in house produced recombinant allergens. The use of frozen PBMC for CytoBas has been validated in previous studies showing that live basophils were readily detectable in thawed PBMC, carrying reliably high levels of surface IgE, and specifically bound to recombinant allergen tetramers (17, 18).

Correlation between serum IgE levels and CytoBas staining intensities for each of the allergenic components only yielded a weak association between the ranks when using Spearman correlation. This might reflect that a biological difference in the measurements: soluble specific-IgE in serum versus specific IgE bound on the surface of basophils. In line with this, previous studies have shown that serum IgE levels do not correlate well with the severity of food allergy (35). As the IgE on immune effector cells is directly involved in the allergic response, the CytoBas assay has the potential to be more informative for determining the sensitization profile and severity of allergic responses (36). Confirming applicability requires future studies in a larger cohort with direct comparison to challenge tests, which are more frequently performed in children than in adults.

Using Ara h 1, Ara h 2, and Ara h 3 tetramers, we effectively identified peanut allergen-specific Bmem populations by flow cytometry. The inclusion of these three major allergens enabled parallel assessment of multiple specificities and revealed significantly higher numbers of allergen-specific Bmem in allergic patients than in controls.

Allergen-specific Bmem showed distinct Ig isotype and IgG subclass expression distributions. Ara h 2-specific Bmem of the peanut cohort displayed a significant increase in the IgE subset compared to non-sensitized controls, which was not detected in Ara h 1- or Ara h 3 specific Bmem (**Fig 5 B**). The IgG1 subclass expression was largely increased in the Ara h 3+ specific Bmem, and a similar trend was detected for Ara h 1 specific Bmem at the expense of the IgM+IgD+ subset. All Ara h -specific Bmem displayed a non-significant reduction in the IgA subset compared to controls (**Fig 5B, C**). These differences may relate to structural and biochemical distinctions between the allergenic proteins as Ara h 1 and Ara h 3 are members of the Cupin superfamily, and Ara h 2 is a member of the Prolamin superfamily, which could influence epitope exposure and subsequent B-cell activation pathways. Another possible explanation would be the varied allergen stability, with Ara h 2 displaying stronger resistance to proteolytic enzymes when digested compared to other allergen family members (37). Furthermore, the balance between helper-T(h) and regulatory T cell (Treg) subsets at time of sensitization could affect the immunological and clinical outcome of peanut allergy (41). Emerging evidence is also pointing to the specific follicular helper-T (TfH) cell milieu during the initial sensitization. The fine equilibrium between TfH_2_ cells, that promote a medium-affinity IgE, and TfH_13_ cells, which are essential for high affinity/ anaphylactic IgE, could dictate the distinct immunoglobulin subclass expansions (42).

We further observed a significantly higher frequency and absolute count of allergen-specific Bmem exhibiting a Type 2 phenotype (CD23 IL-4Rα) in the peanut allergic cohort, consistent with previous reports in allergic asthma, atopic dermatitis and peanut allergy (21, 38). This subset represents a reservoir for pathogenic IgE, as type 2 Bmem have been shown to express germline *IGHE* transcripts (21), and can differentiate into IgE-producing plasma cells upon allergen re-exposure (20). Thus, type 2 Bmem play a central role in maintaining IgE memory and could serve as a potential biomarker to monitor the effects of AIT. We recently reported an initial expansion of type 2 Bmem in patients undergoing AIT for house dust mite allergy, which contracts at 18 months, indicating an initial expansion before retraction (39). We further observed that in early stages of AIT for ryegrass pollen or bee venom allergy, type 2 Bmem from allergic individuals expressed more frequently integrin β1 (CD29) and/or IgG4 (19, 37). In the current single sampling study, no significant differences in these markers were detected. However, these can change in a long-term AIT regimen. CD29 is involved in regulating BCR signalling and the upregulation may inhibit B cell response upon re-exposure (40), which could potentially contribute to immunological desensitization on treatment to the specific allergen.

In conclusion, we have demonstrated the utility of recombinant peanut allergen components in CytoBas for component-resolved diagnosis and risk stratification of patients with peanut sensitization. Furthermore, the peanut allergen components can be used to evaluate the allergen-specific Bmem compartment in peanut allergic individuals enabling immune monitoring of the pathogenic Type 2 memory B-cell subset over the course of peanut AIT as a companion diagnostic.

## Acknowledgments

We thank the staff of ARAFlowCore (Monash University) for flow cytometry support, and Dr. Bruce Wines (Burnet Institute) for support with recombinant protein production.

## Author contributions

Conceptualization: MCvZ, MH, REOH

Methodology: MCvZ, REOH, PMH, SR

Investigation: SR, PMA, RF, KD

Funding acquisition: MCvZ, MH, REOH

Supervision: MCvZ, REOH

Writing – original draft: SR, MCvZ

Writing – review & editing: RF, KD, PMH, MH, REOH

## Data Availability Statement

The data that support the findings of this study are available from the corresponding author upon reasonable request.

## Abbreviations

Ara h: Arachis hypogaea
AIT: allergen immunotherapy
AUC: area under the curve
BAT: basophil activation test
Bmem: memory B cell
CRD: component resolved diagnostics
IgE: Immunoglobulin E
ImmunoCAP: molecular diagnostic test for allergen-specific IgE in blood
MOI: multiplicity of infection
PBMC: peripheral blood mononuclear cells
ROC: Receiver Operating Characteristic
SPT: skin prick test
Treg: regulatory T cell
TfH: T follicular helper cell

## REFERENCES

1. Abrams EM, Chan ES, Sicherer S. Peanut Allergy: New Advances and Ongoing Controversies. Pediatrics. 2020;145(5).

2. Upton JEM, Toscano-Rivero D, Ke D, Berenjy A, Lejtenyi D, Beaudette L, et al. Peanut Oral Immunotherapy Using 30 and 300 mg Maintenance Doses. J Allergy Clin Immunol Pract. 2026;14(1):223–32.e7.

3. Reinwald S, Rolland JM, O’Hehir RE, van Zelm MC. Peanut oral immunotherapy: current trends in clinical trials. Immunother Adv. 2022;2(1):ltac004.

4. Nagy NA, Schnoegl D, Houghton V, O’Mahony L, Santos AF, Knol EF, et al. Sequence of Immunological Events During IgE-Mediated Allergic Reactions to Food. Allergy. 2025.

5. Kaur N, Mehr S, Katelaris C, Wainstein B, Altavilla B, Saad R, et al. Added Diagnostic Value of Peanut Component Testing: A Cross-Sectional Study in Australian Children. J Allergy Clin Immunol Pract. 2021;9(1):245–53.e4.

6. Nilsson C, Berthold M, Mascialino B, Orme ME, Sjolander S, Hamilton RG. Accuracy of component-resolved diagnostics in peanut allergy: Systematic literature review and meta-analysis. Pediatr Allergy Immunol. 2020;31(3):303–14.

7. Polomska J, Dydak P, Sozanska B, Sikorska-Szaflik H. Peanut Allergy and Component-Resolved Diagnostics Possibilities-What Are the Benefits? Nutrients. 2023;15(24).

8. Kukkonen AK, Pelkonen AS, Makinen-Kiljunen S, Voutilainen H, Makela MJ. Ara h 2 and Ara 6 are the best predictors of severe peanut allergy: a double-blind placebo-controlled study. Allergy. 2015;70(10):1239–45.

9. Dang TD, Tang M, Choo S, Licciardi PV, Koplin JJ, Martin PE, et al. Increasing the accuracy of peanut allergy diagnosis by using Ara h 2. Journal of Allergy and Clinical Immunology. 2012;129(4):1056–63.

10. Cabanillas B, Jappe U, Novak N. Allergy to Peanut, Soybean, and Other Legumes: Recent Advances in Allergen Characterization, Stability to Processing and IgE Cross-Reactivity. Mol Nutr Food Res. 2018;62(1).

11. Mak R, Chan ES, Irvine M, Huang JY, Hethey J, Hartstein S, et al. Comparing approaches to ordering peanut component-resolved diagnostics to reduce the need for oral food challenges. J Allergy Clin Immunol Glob. 2025;4(2):100440.

12. Santos AF, Du Toit G, O’Rourke C, Becares N, Couto-Francisco N, Radulovic S, et al. Biomarkers of severity and threshold of allergic reactions during oral peanut challenges. J Allergy Clin Immunol. 2020;146(2):344–55.

13. Calvani M, Bianchi A, Reginelli C, Peresso M, Testa A. Oral Food Challenge. Medicina (Kaunas). 2019;55(10).

14. Knol EF. Requirements for effective IgE cross-linking on mast cells and basophils. Mol Nutr Food Res. 2006;50(7):620–4.

15. Sonder SU, Plassmeyer M, Loizou D, Alpan O. Towards standardizing basophil identification by flow cytometry. Front Allergy. 2023;4:1133378.

16. Santos AF, Alpan O, Hoffmann H-J. Basophil activation test: Mechanisms and considerations for use in clinical trials and clinical practice. Allergy. 2021;76(8):2420–32.

17. McKenzie CI, Varese N, Aui PM, Wines BD, Hogarth PM, Thien F, et al. CytoBas: Precision component-resolved diagnostics for allergy using flow cytometric staining of basophils with recombinant allergen tetramers. Allergy. 2021;76(10):3028–40.

18. Hsin L, Varese N, Aui PM, Wines BD, von Borstel A, Mascarell L, et al. Accurate determination of house dust mite sensitization in asthma and allergic rhinitis through cytometric detection of Der p 1 and Der p 2 binding on Basophils (CytoBas). Journal of Allergy and Clinical Immunology. 2024.

19. McKenzie CI, Varese N, Aui PM, Reinwald S, Wines BD, Hogarth PM, et al. RNA sequencing of single allergen-specific memory B cells after grass pollen immunotherapy: Two unique cell fates and CD29 as a biomarker for treatment effect. Allergy. 2023;78(3):822–35.

20. Koenig JFE, Knudsen NPH, Phelps A, Bruton K, Hoof I, Lund G, et al. Type 2-polarized memory B cells hold allergen-specific IgE memory. Sci Transl Med. 2024;16(733):eadi0944.

21. Ota M, Hoehn KB, Fernandes-Braga W, Ota T, Aranda CJ, Friedman S, et al. CD23(+)IgG1(+) memory B cells are poised to switch to pathogenic IgE production in food allergy. Sci Transl Med. 2024;16(733):eadi0673.

22. von Borstel A, Reinwald S, Aui PM, McKenzie CI, Varese N, Hogarth PM, et al. Expansion of phenotypically modified type 2 memory B cells after allergen immunotherapy. Allergy. 2025;80(3):867–9.

23. Chan SK, Pomés A, Hilger C, Davies JM, Mueller G, Kuehn A, et al. Keeping Allergen Names Clear and Defined. Front Immunol. 2019;10:2600.

24. Hartley GE, Edwards ESJ, Aui PM, Varese N, Stojanovic S, McMahon J, et al. Rapid generation of durable B cell memory to SARS-CoV-2 spike and nucleocapsid proteins in COVID-19 and convalescence. Science Immunology. 2020;5(54):eabf8891.

25. Tani H, Nishijima M, Ushijima H, Miyamura T, Matsuura Y. Characterization of cell-surface determinants important for baculovirus infection. Virology. 2001;279(1):343–53.

26. Arakawa T, Timasheff SN. Mechanism of protein salting in and salting out by divalent cation salts: balance between hydration and salt binding. Biochemistry. 1984;23(25):5912–23.

27. Schatz PJ. Use of peptide libraries to map the substrate specificity of a peptide-modifying enzyme: a 13 residue consensus peptide specifies biotinylation in Escherichia coli. Bio/technology. 1993;11(10):1138–43.

28. Fryer HA, Hartley GE, Edwards ES, O’Hehir RE, van Zelm MC. Humoral immunity and B-cell memory in response to SARS-CoV-2 infection and vaccination. Biochemical Society Transactions. 2022;50(6):1643–58.

29. Edwards JM, Andrews MC, Burridge H, Smith R, Owens C, Edinger M, et al. Design, optimisation and standardisation of a high-dimensional spectral flow cytometry workflow assessing T-cell immunophenotype in patients with melanoma. Clin Transl Immunology. 2023;12(9):e1466.

30. van Zelm MC, McKenzie CI, Varese N, Rolland JM, O’Hehir RE. Recent developments and highlights in immune monitoring of allergen immunotherapy. Allergy. 2019;74(12):2342–54.

31. Ojaniemi I, Salmivesi S, Tikkakoski A, Karjalainen J, Lehtimäki L, Schultz R. Are peanut oral food challenges still useful? An evaluation of children with suspected peanut allergy, sensitization to Ara h 2 and controlled asthma. Allergy, Asthma & Clinical Immunology. 2022;18(1):100.

32. van Zelm MC, McKenzie CI, Varese N, Rolland JM, O’Hehir RE. Advances in allergen-specific immune cell measurements for improved detection of allergic sensitization and immunotherapy responses. Allergy. 2021;76(11):3374–82.

33. van Zelm MC, O’Hehir RE, McKenzie CI. A recent patent in allergy & immunology: Biomarkers on allergen-specific memory B cells to predict allergen immunotherapy outcome. Allergy. 2024;79(8):2295–7.

34. Keet C, Plesa M, Szelag D, Shreffler W, Wood R, Dunlop J, et al. Ara h 2-specific IgE is superior to whole peanut extract-based serology or skin prick test for diagnosis of peanut allergy in infancy. J Allergy Clin Immunol. 2021;147(3):977–83.e2.

35. Ponce M, Diesner SC, Szépfalusi Z, Eiwegger T. Markers of tolerance development to food allergens. Allergy. 2016;71(10):1393–404.

36. Dölle-Bierke S, Höfer V, Francuzik W, Näher A-F, Bilo MB, Cichocka-Jarosz E, et al. Food-Induced Anaphylaxis: Data From the European Anaphylaxis Registry. The Journal of Allergy and Clinical Immunology: In Practice. 2023;11(7):2069–79.e7.

37. McKenzie CI, Reinwald S, Averso B, Spurrier B, Satz A, von Borstel A, et al. Subcutaneous immunotherapy for bee venom allergy induces epitope spreading and immunophenotypic changes in allergen-specific memory B cells. J Allergy Clin Immunol. 2024;154(6):1511–22.

38. Aranda CJ, Gonzalez-Kozlova E, Saunders SP, Fernandes-Braga W, Ota M, Narayanan S, et al. IgG memory B cells expressing IL4R and FCER2 are associated with atopic diseases. Allergy. 2023;78(3):752–66.

39. Hsin L, Reinwald S, von Borstel A, Aui PM, Deckert K, Hogarth PM, et al. Biphasic Effects on Allergen-Specific Type 2 Memory B Cells Over 18 Months Sublingual Immunotherapy for House Dust Mite Allergy. Allergy. 2026;81(6):2172–83.

40. Andreani V, Ramamoorthy S, Fässler R, Grosschedl R. Integrin β1 regulates marginal zone B cell differentiation and PI3K signaling. J Exp Med. 2023;220(1).

41. Turcanu V, Brough HA, Du Toit G, Foong RX, Marrs T, Santos AF, Lack G. Immune mechanisms of food allergy and its prevention by early intervention. Curr Opin Immunol. 2017 Oct;48:92–98.

42. Lama JK, Kita H. T_FH_ cells regulate antibody affinity and determine the outcomes of anaphylaxis. J Allergy Clin Immunol. 2022 Nov;150(5):1042–1044

