## Supplemental Tables and Figures for "Detection of Peanut Allergy Sensitization and Monitoring of Type 2 Memory B cells Using Recombinant Peanut Allergen Components"

**Supplementary Tables** (n=6)

**Supplementary Table 1. Demographics of patient allergic patients.**

| **Peanut allergic individual** | **sex** | **Age**  **(year)** | **Total serum IgE (kU/L)** | **Peanut specific serum IgE (kU/L)** | **RAST category**  **(1-5)** | **Grass pollen allergy y/n** |
| --- | --- | --- | --- | --- | --- | --- |
| 1 | M | 18-25 | 663 | >100 | 5 | y |
| 2 | M | 18-25 | 94 | 4.26 | 3 | not tested |
| 3 | M | 18-25 | 208 | 88.2 | 5 | y |
| 4 | M | 18-25 | 378 | 23.3 | 4 | y |
| 5 | M | 18-25 | 91 | 44.2 | 4 | not tested |
| 6 | M | 18-25 | 106 | 2.61 | 2 | y |
| 7 | F | 18-25 | 290 | 59.4 | 5 | n |
| 8 | F | 18-25 | 583 | 3.81 | 3 | y |
| 9 | M | 18-25 | 250 | 1.78 | 2 | y |
| 10 | M | 26-30 | 1689 | 46.6 | 4 | y |
| 11 | M | 26-30 | 289 | 0.65 | 1 | y |
| 12 | F | 26-30 | 309 | 2.03 | 2 | not tested |
| 13 | M | 26-30 | 144 | 40.4 | 4 | not tested |
| 14 | F | 26-30 | *NT* | 0.98 | 2 | y |
| 15 | F | 26-30 | 939 | 15.7 | 3 | n |
| 16 | M | 26-30 | 4580 | 26 | 4 | y |
| 17 | M | 26-30 | 213 | 1.93 | 2 | y |
| 18 | M | 26-30 | 277 | 80.1 | 5 | y |
| 19 | F | 26-30 | 72 | 0.35 | 1 | y |
| 20 | F | 31-35 | 160 | 46.8 | 4 | n |
| 21 | F | 31-35 | 1114 | 8.4 | 2 | y |
| 22 | M | 31-35 | 360 | 13.2 | 3 | not tested |
| 23 | F | 31-35 | 74 | 10.2 | 3 | n |
| 24 | M | 31-35 | 1420 | 9.37 | 3 | y |
| 25 | M | 36-40 | 5450 | 10.6 | 3 | y |
| 26 | F | 41-45 | 98 | 30.7 | 4 | not tested |
| 27 | F | 41-45 | 29 | 1.85 | 2 | n |

**Supplementary Table 2. Demographics of non-peanut sensitized controls.**

| **non-peanut sensitized**  **individual** | **sex** | **Age at analysis** |
| --- | --- | --- |
| 1 | F | 26-30 |
| 2 | F | 26-30 |
| 3 | F | 26-30 |
| 4 | M | 26-30 |
| 5 | M | 31-35 |
| 6 | F | 31-35 |
| 7 | M | 31-35 |
| 8 | M | 31-35 |
| 9 | M | 36-40 |
| 10 | F | 36-40 |
| 11 | M | 41-45 |
| 12 | M | 41-45 |
| 13 | F | 46-50 |
| 14 | F | 46-50 |
| 15 | M | 61-65 |

**Supplementary Table 3. List of antibodies used in TruCount flow cytometry**

| **Marker** | **Fluorochrome** | **Clone** | **Source** | **Cat. number** | **Vol per test (µL/70 µL)** | **Tube *** |
| --- | --- | --- | --- | --- | --- | --- |
| **CD3** | FITC | UCHT1 | BD Biosciences | 555332 | 3 | 1 |
| **CD4** | PE-Cy7 | SFCI12T4D11 | Beckman Coulter | 6607101 | 0.2 | 1 |
| **CD8A** | APC-Cy7 | SK1 | BD Biosciences | 560179 | 4 | 1 |
| **CD16** | PE | B73.1 | BioLegend | 360704 | 0.5 | 1 |
| **CD19** | APC | SJ25C1 | BioLegend | 561742 | 0.4 | 1 |
| **CD45** | PerCP-Cy5.5 | 2D1 | BD Biosciences | 340953 | 5 | 1 |
| **CD56** | PE | B159 | BD Biosciences | 555516 | 5 | 1 |

**Supplementary Table 4. List of antibodies used in Cytobas flow cytometry**

| **Allergen tetramer** | **Reagent** | **Fluorochrome** | **Clone** | **Source** | **Cat. number** | **Vol per test (µL)** |
| --- | --- | --- | --- | --- | --- | --- |
| **[Ara h 1]**_4_ | Streptavidin | BUV395 | - | BD Biosciences | 564176 | 6.2 |
| **[Ara h 2]**_4_ | Streptavidin | PE | - | Life Technologies | S866 | 5.9 |
| **[Ara h 3]**_4_ | Streptavidin | BUV615 | - | BD Biosciences | 613013 | 3.7 |
| **[Ara h 6]**_4_ | Streptavidin | BV480 | - | BD Biosciences | 564876 | 0.8 |
| **[Ara h 8]**_4_ | Streptavidin | BUV737 | - | BD Biosciences | 564293 | 6.2 |
| **[Ara h 9]**_4_ | Streptavidin | BV711 | - | BD Biosciences | 563262 | 3.8 |
|  | CD45 | PerCP-Cy5.5 | 2D1 | BD Biosciences | 340953 | 2 |
|  | CD123 | cFluorBYG781 | SK3 | Cytek Biosciences | R7-11004 | 0.1 |
|  | IgE | FITC | Goat anti-human polyclonal | Life Technologies | H15701 | 1.25 |
|  | HLA-DR | APC | L243 | BioLegend | 307610 | 1 |
|  | Viability Dye | viaDye Red | - | Cytek Biosciences | R7-60008 | 0.0025 |

**Supplementary Table 5. List of antibodies used in memory B cell flow cytometry**

| **Marker** | **Fluorochrome** | **Clone** | **Source** | **Cat. number** | **Vol per test (µL/250 µL)** |
| --- | --- | --- | --- | --- | --- |
| **CD3** | BUV805 | UCHT1 | BD Biosciences | 612895 | 6.25 |
| **CD19** | BUV563 | SJ25C1 | BD Biosciences | 612917 | 1.5 |
| **CD21** | BV711 | B-ly4 | BD Biosciences | 563163 | 1.25 |
| **CD23** | APC-Cy7 | EBVCS-5 | Biolegend | 338520 | 2.5 |
| **CD27** | cFluor BYG781 | O323 | Cytek | Custom | 1.25 |
| **CD29** | cFluor BYG750 | MEM-101A | Cytek | Custom | 1.25 |
| **CD38** | cFluor V610 | HB7 | Cytek | Custom | 6.25 |
| **CD45** | cFluor V547 | HI30 | Cytek | R7-20011 | 1.25 |
| **CD124 (IL4Rα)** | PE | G9077F6 | Biolegend | 355004 | 1.66 |
| **IgA** | cFluorB515 | HP6123 | Cytek | Custom | 3.125 |
| **IgD** | BB700 | IA6-2 | BD Biosciences | 566538 | 2.5 |
| **IgE** | cFluor R840 | HP6123 | Cytek | Custom | 2.5 |
| **IgG1** | cFluor B532 | 12G8G11 | Cytek | Custom | 1.56 |
| **IgG2** | cFluor R685 | HP6002 | Cytek | Custom | 3.125 |
| **IgG3** | cFluor R668 | HP6047 | Cytek | Custom | 3.125 |
| **IgG4** | APC | SAG4 | Cytognos | CYT-IGG4AP | 5 |
| **IgM** | cFluor V450 | MHM-88 | Cytek | Custom | 0.625 |
| **[Ara h 1]_4_ Strep** | BV480 | - | BD Biosciences | 564876 |  |
| **[Ara h 1]_4_ Strep** | BUV737 | - | BD Biosciences | 564293 |  |
| **[Ara h 2]_4_ Strep** | BUV395 | - | BD Biosciences | 564176 |  |
| **[Ara h 2]_4_ Strep** | BV421 | - | BioLegend | 405225 |  |
| **[Ara h 3]_4_ Strep** | BUV496 | - | BD Biosciences | 564666 |  |
| **[Ara h 3]_4_ Strep** | BV650 | - | Biolegend | 405232 |  |
| **Viability dye** | Live Dead Blue | - | Life Technologies | L23105 | 6.25 (1:40 dil) |

**Supplementary Table 6. List of MFI ratio of allergen/strep-only per patient per allergen component.**

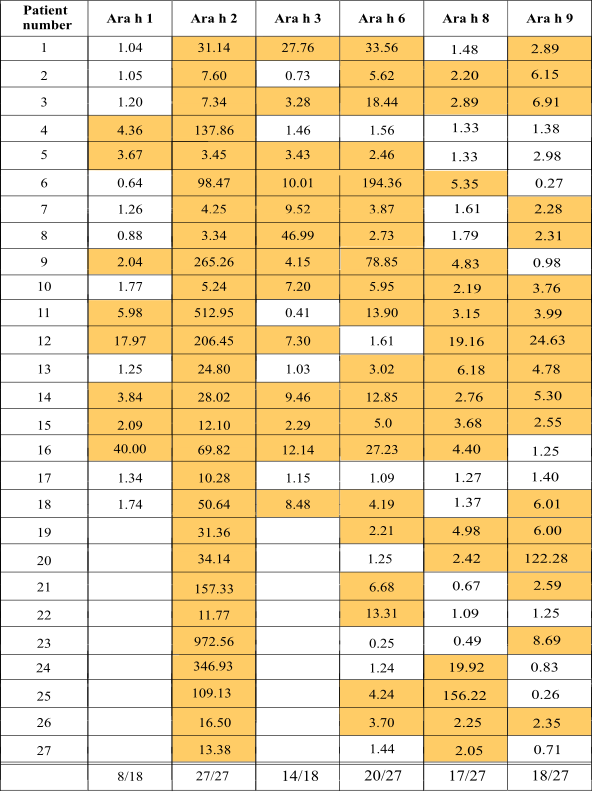

Ratio of or above 2 are indicated in yellow, numbers of patients positive of the total cohort noted below.

**Supplementary Figures** (n=4)

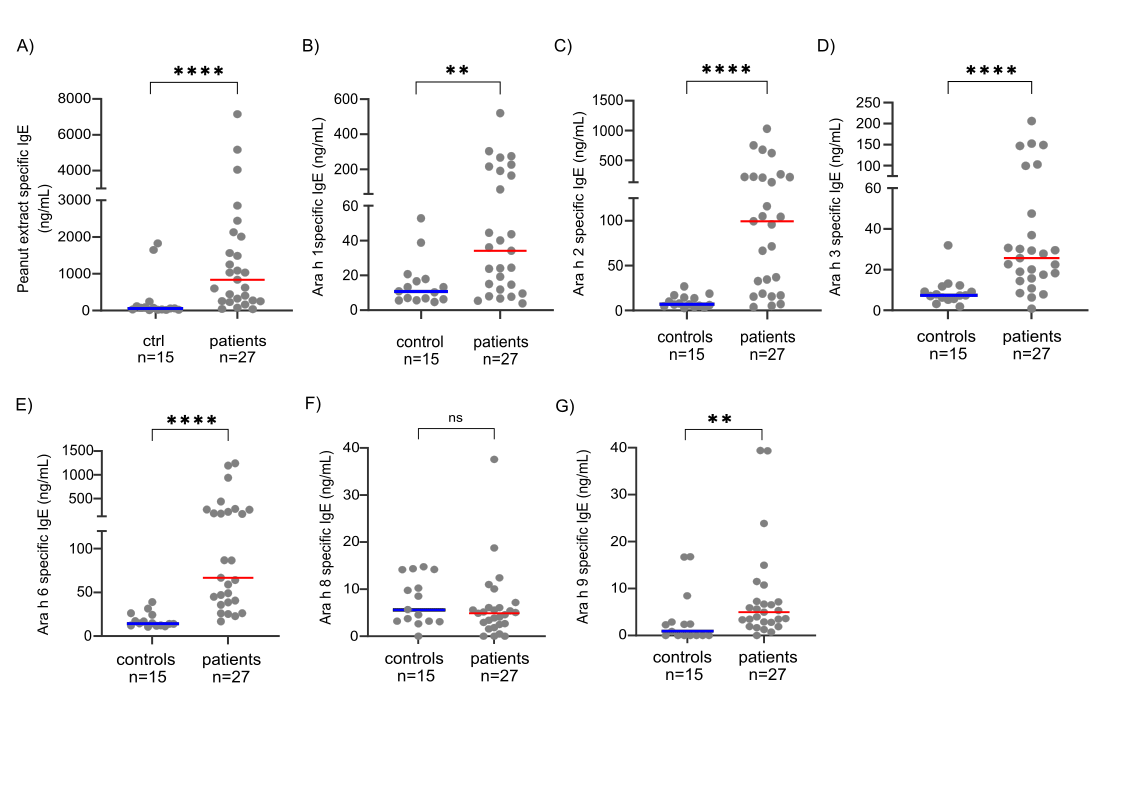

**Supplementary Figure 1. Serum peanut-specific IgE in non-sensitized controls and peanut-sensitized patients.** 15 non-peanut sensitized controls and 27 peanut-sensitized patients were tested for serum IgE to (**A**) peanut extract, **(B)** Ara h 1, **(C)** Ara h 2, **(D)** Ara h 3, **(E)** Ara h 6, **(F)** Ara h 8 and **(G)** Ara h 9 using ELISA. Statistics: Mann-Whitney non-parametric test ** p=0.01 *** p=0.001 **** p=0.0001.

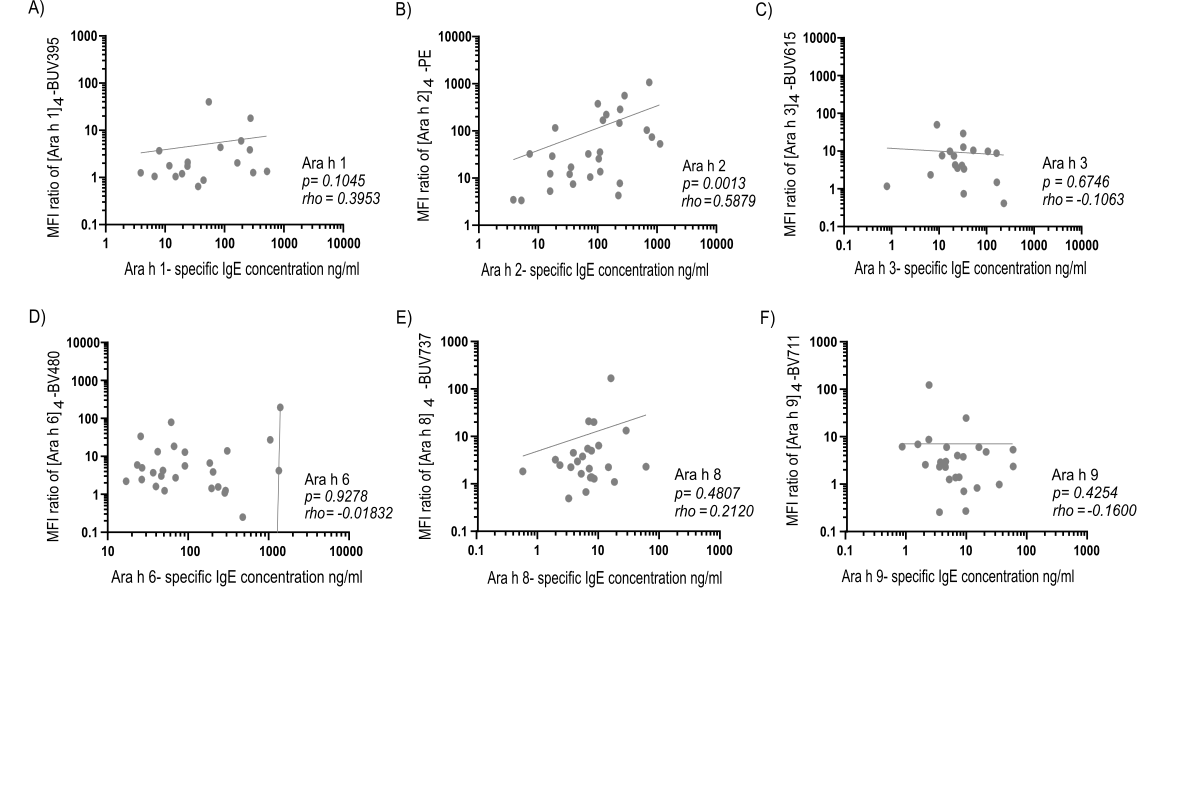
**Supplementary Figure 2.** **Correlations between specific IgE binding intensities on basophils and serum-specific IgE concentrations.** Correlation plots between circulating specific IgE levels and basophil tetramer binding staining intensities for all assessed allergic individuals for (**A**) Ara h 1, **(B)** Ara h 2, **(C)** Ara h 3, **(D)** Ara h 6, **(E)** Ara h 8, and **(F)** Ara h 9. Individual data points are shown with log-log regression lines. Spearman's rank correlation tests were conducted, the p-values indicate the significance of the correlation tests.

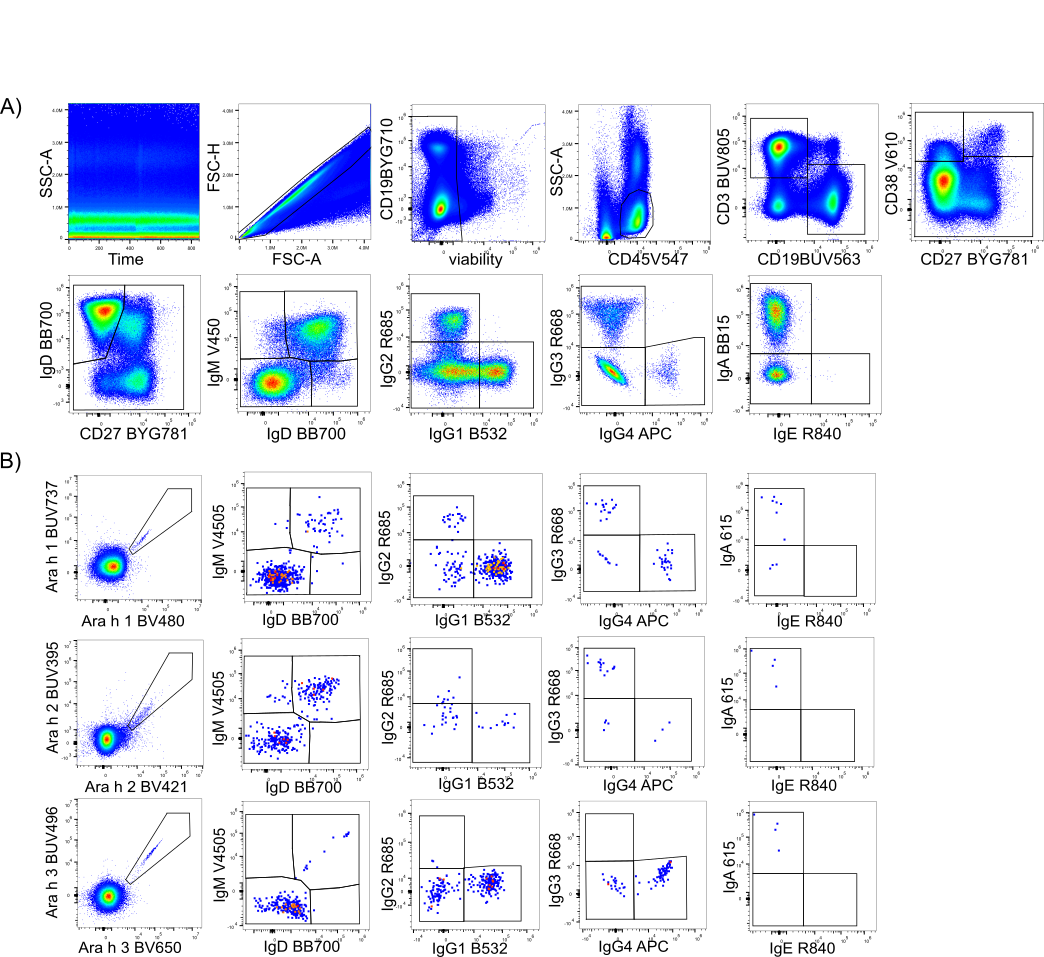

**Supplementary Figure 3. Allergen-specific Bmem gating strategy.** (**A**) From all events, the singlet live cells were gated with subsequent gating of the lymphocyte population using CD45 and SSC. T (CD3+) and B (CD19+) cells were discriminated within lymphocytes, followed by the subsetting of B cells into CD38^dim^ mature B cells, transitional B cells and plasmablasts. Within the CD38dim cells, memory B cells (Bmem) were defined by excluding the naive B population (CD27-IgD+). Bmem were separated using IgM and IgD into unswitched Bmem (IgM+only, IgD+ only and IgM+IgD+) and Immunoglobulin (Ig) class-switched populations. Ig-switched subsets were then defined by the differential expression of IgG1, IgG2, IgG3, IgG4, IgA and IgE. (**B**) Ara h 1, 2, or 3 specific Bmem were separated using IgM and IgD into unswitched Bmem and Ig class switched populations as done in total Bmem. Ig-switched subsets were defined by the differential expression of IgG1, IgG2, IgG 3, IgG 4, IgA and IgE.

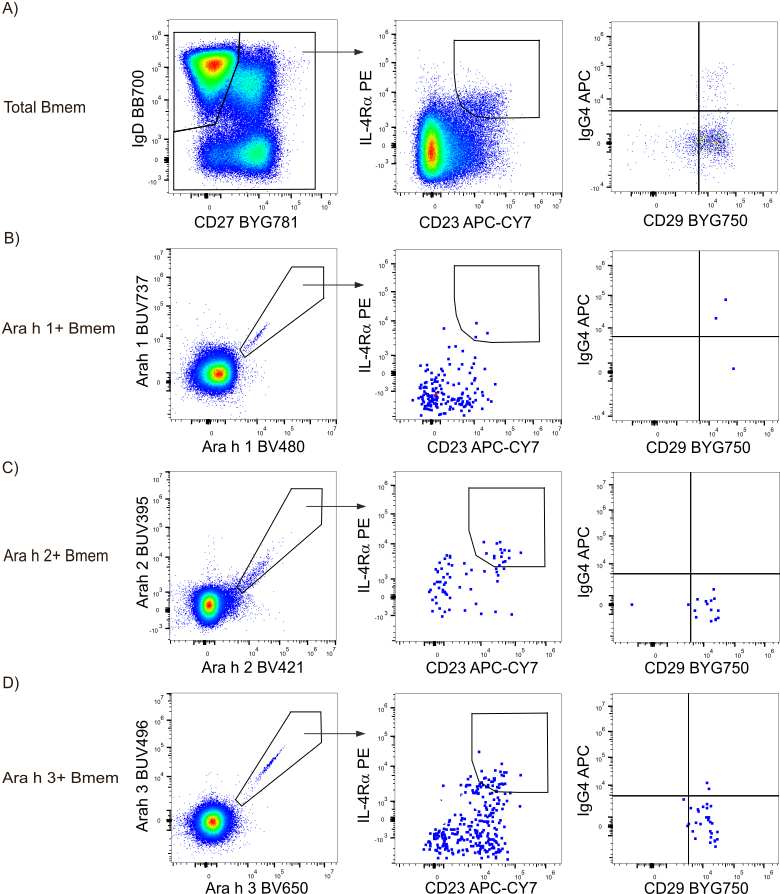

**Supplementary Figure 4. Type 2 memory B cell gating strategy.** Type 2 memory B cells were defined by co-expression of CD23 and IL-4Ra within total **(A)** or Ara h 1+ specific **(B)**, Ara h 2+ specific **(C)** or Ara h 3+ specific memory B cells **(D).** Within the type 2 Bmem subset, IgG4 and CD29 expression were determined.
